# It’s All in the Details: Guiding Fine-Feature Characteristics in Artificial Medical Images using Diffusion Models

**DOI:** 10.1101/2025.05.01.25326784

**Authors:** Elfi I.S. Hofmeijer, Xinrui Zu, Ferdi van der Heijden, Can Ozan Tan

## Abstract

We sought to develop a diffusion model-based framework that guides both larger anatomical structures and fine features to generate radiographic images that accurately reflect pathological characteristics.

The model is based on a latent diffusion model that is extended to include coarse- and fine-feature guidance. The feedback of an independent classifier network, trained to identify malignant features, was used to provide the fine-feature guidance. We compared the accuracy of this model to that attained by one without fine-feature guidance and by a standard generative adversarial network. We used the area under ROC to compare accuracy across the networks in representing malignant features of lung nodules and gliomas on 44,924 lung CT and 6,376 MRI 2D images (annotated by trained radiologists). Statistical significance was assessed using bootstrapped p-values.

For each dataset, the model generated artificial images comparable to original ones. Benign vs malignant classification accuracy without fine-feature guidance was 70% (CT), 81% (MRI). Fine-feature guidance increased the accuracy to 85.5%, 86%, for, respectively, CT and MRI images (vs unguided, *p* < 0.001, *p* < 0.001).

It is feasible to use independent classifier guidance to create artificial radiographic images that accurately reflect fine features across pathologies and imaging modalities.

## Introduction

There has been a growing emphasis on the utility of generative models in the future of medicine^1^ ranging from the use of electronic health records to predict patient outcomes ^2^ to drug development ^3^. Most of these models present considerable advances toward creating virtual representations of actual cases in the healthcare setting. One critical component is the generation of radiologic images across multiple modalities (e.g., CT, x-ray, MRI) that reflect detailed radiographic representation of multiple pathologies. Radiologic images are one of the most common components of clinical diagnosis, prognosis, and progress.^4^ Thus, the ability to generate artificial images that reflect clinical features across different modalities is a key technology that can enable “simulation” of presentation, diagnoses, prognosis, and trajectories at the level of individual patients.^5,6^

Several lines of research have shown that artificial radiographic images can be generated across different imaging modalities ^7,8^ and body parts ^8,9^. We have previously shown that these images can be reasonably realistic to an extent where overall representation and features of generated images can be indistinguishable by trained radiologists^10^. However, in addition to overall features, virtual representation of radiographic images must also reflect fine pathological features accurately, which remains elusive ^9,11^.

We sought to investigate if we can improve representation of fine features along with coarse ones. Our goal is to represent large anatomical features as well as detailed features reflective of pathological characteristics evident in actual radiographic images. We demonstrate that the used architecture is able to generate clinical images across two main modalities (CT and MRI), body parts (lungs and brain), and pathologies (malignant vs benign lung nodules and brain tumor).

## Materials and Methods

The basic architecture of the created model, “Latent Diffusion for Fine Feature Guidance (DiFine)”, is based on the Latent Diffusion Models (LDMs) ^12^, but includes an independent classifier network within the model. This independent classifier is able to learn fine discriminating features, the characteristics of an object as a certain class, that are used to guide the diffusion model. In particular, we use (1) a semantic label map to define the location and size of the pathological area as well as other coarse features and (2) a trained independent classifier network to provide more detailed information on sub-types of the pathology.

### Network Architecture

The basis of the DiFine model is the LDM framework and its implementation ^13^. We chose to use an autoencoder based on a residual neural network ^14^ trained to learn a reasonable representation of the 128×128×3 latent space of an 512×512×1 image. The choice of latent space size is arbitrary and based on the trade-off between information loss and computation gain.

To guide the representation of the artificial radiographic image, we implemented both coarse- and fine-feature guidance based on previous work^15,16^ in tandem. In contrast to the prior work, DiFine is a unique combination of the two guidance models, and the fine-feature guidance is implemented in an *in-painting style* manner as opposed to the class guidance on full images that is a common feature of prior models. This in-painting style reflects our focus on a given area of interest within the artificial image. This area is cropped and used to train an independent classifier. This results in an *in-painted* image of the area of interest (Figure 1). The network details, specifically the coarse- and fine-feature guidance, are described in more detail in the Supplemental Material. All code can be found at https://github.com/UT-RAM-AIM/FineFeature-guidance.

**Figure 1.**
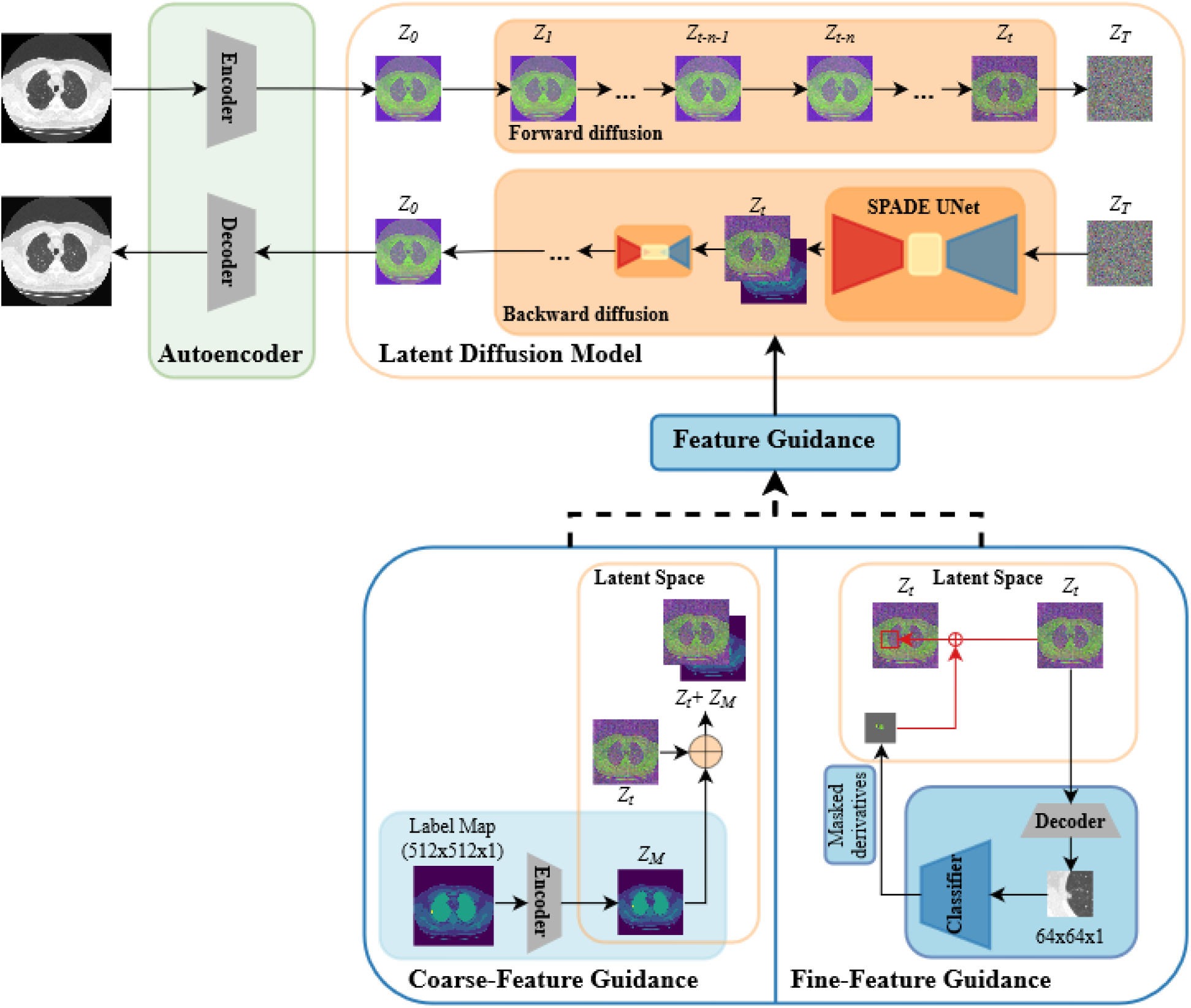
DiFine architecture; the original image is encoded into the latent space and noise is added through the forward diffusion process resulting in a noisy image. This noisy image is denoised during the backward diffusion process and decoded into original pixel space to obtain an artificial image. Various (de-)noising steps are depicted, including the denoising UNet with SPADE blocks. The coarse-feature guidance representing the semantic label map is encoded into the latent space and concatenated with the noisy image at that time step. The fine-feature guidance represents the pathological area cropped out of the noisy images, and the masked information obtained from the classifier is in-painted in the noisy image.

### Datasets and Image Preprocessing

We used the Lung Image Database Consortium (LIDC) and Image Database Resource Initiative (IDRI) database with 805 low-dose 3D chest CT scans^17,18^ and the international multimodal Brain Tumor Segmentation Challenge (BraTS2019) dataset with 335 3D head MRI scans^19–23^. Both datasets are anonymized and are publicly available for general research. The first dataset was obtained through The Cancer Imaging Archive ^24^ and included 512x512 slices along with their pathological classification and annotation of objects (malignant vs benign nodules, for fine-feature guidance). This is similar for the second dataset except slices are 240×240. The semantic label maps for coarse-feature guidance were obtained algorithmically (further described in the Supplemental Material). A summary of modality and body part, semantic label map labels, and relevant pathological classification are summarized Table 1. All datasets are split on a case level into a train- and test set (Table 1). Further details of the preprocessing steps for each dataset are described in the Supplemental Material.

**Table 1.** Summary of the course features present in the two datasets: modality and organ, semantic labels indicated in the semantic label map, number of 2D images used as training and test set, and total number of unique scans.

| Dataset | Modality | Semantic labels in map | Number of slices in the training/test sets | Number of unique scans |
| --- | --- | --- | --- | --- |
| **LIDC-IDRI** <sup>18</sup> | Lung CT | 1. Background<br>2. Body<br>3. Low-dense tissue<br>4. High-dense tissue<br>5. Lung<br>6. Lung nodule | 39,803 / 5,121 | 805 |
| **BRATS2019** <sub>22,23</sub> | Brain MRI | 1. Background<br>2. White matter<br>3. Grey matter<br>4. Necrotic core<br>5. GD-enhanced core | 5,228 / 1,148 | 335 |

Given its clinical relevance, we focused on the malignancy of the main pathology to guide fine features. We chose only the main pathology as these were already annotated by multiple domain experts. The LIDC-IDRI dataset provides a score for each lung nodule on a scale of 1 (benign) to 5 (malignant) assessed by four experienced chest radiologists. We simplified these scores to obtain a binary designation, where a score < 3 was considered benign, and a score of => 3 was considered malignant. In case of the BraTS2019 dataset, the gliomas were labelled as high- or low-grade and we used these designations without any modification.

### Model Evaluation

By design, the fine features of the pathology are assumed to be the main driver of the type of pathology (benign vs malignant, or high vs. low grade glioblastoma). This is a reasonable assumption given the clinical context. Therefore, comparison between types of pathology (predicted by the independent classifier) on real versus artificial images provides a measure of accuracy in generating fine features on the image. This is different than image-to-image comparison (e.g., dice score) as the latter simply provides a measure of overall pixel-wise accuracy and not of fine features themselves. In this context, while relevant, a high absolute accuracy of the classifier network is not critical; the focus is on creating fine features in an *artificial* image, and not on assessing whether or not features embedded in a *real* image can predict the actual type of pathology. That is, we sought to investigate if the DiFine model is able to encapsulate fine features appropriately using its independent classifier network, and not to achieve the highest accuracy for the latter network.

### Statistics

We derived the receiver operating characteristic (ROC) curves as a measure of accuracy of the DiFine model with and without fine-feature guidance. We used a pairwise bootstrap test ^25,26^ with 10000 bootstrap permutations to compare ROC curves. To compare performance of the DiFine model to predecessors, we further assessed the accuracy of its created artificial images to those created by a trained Generative Adversarial Network (GAN)-based model with SPADE ^27^. We additionally assess the utility of the independent classifier on representing the fine features of the opposite class. To that end, we compared the independent classifier prediction of the created images with the actual class pathology (e.g., benign nodule) to the predictions when the same images created by the DiFine model are guided by the opposite class (e.g., malignant nodule). This provided a sensitivity analysis and an additional validation of the generative model performance.

## Results

Artificial images created by the DiFine model were visually comparable to those of the original images for both datasets (Figure 2). As expected, coarse structures (e.g., lung area, or white matter) in the artificial images adhered to the semantic label maps. The fine-feature guidance reflected visual changes in the pathology area (lung nodules in chest CT as an example, Figure 3).

**Figure 2.**
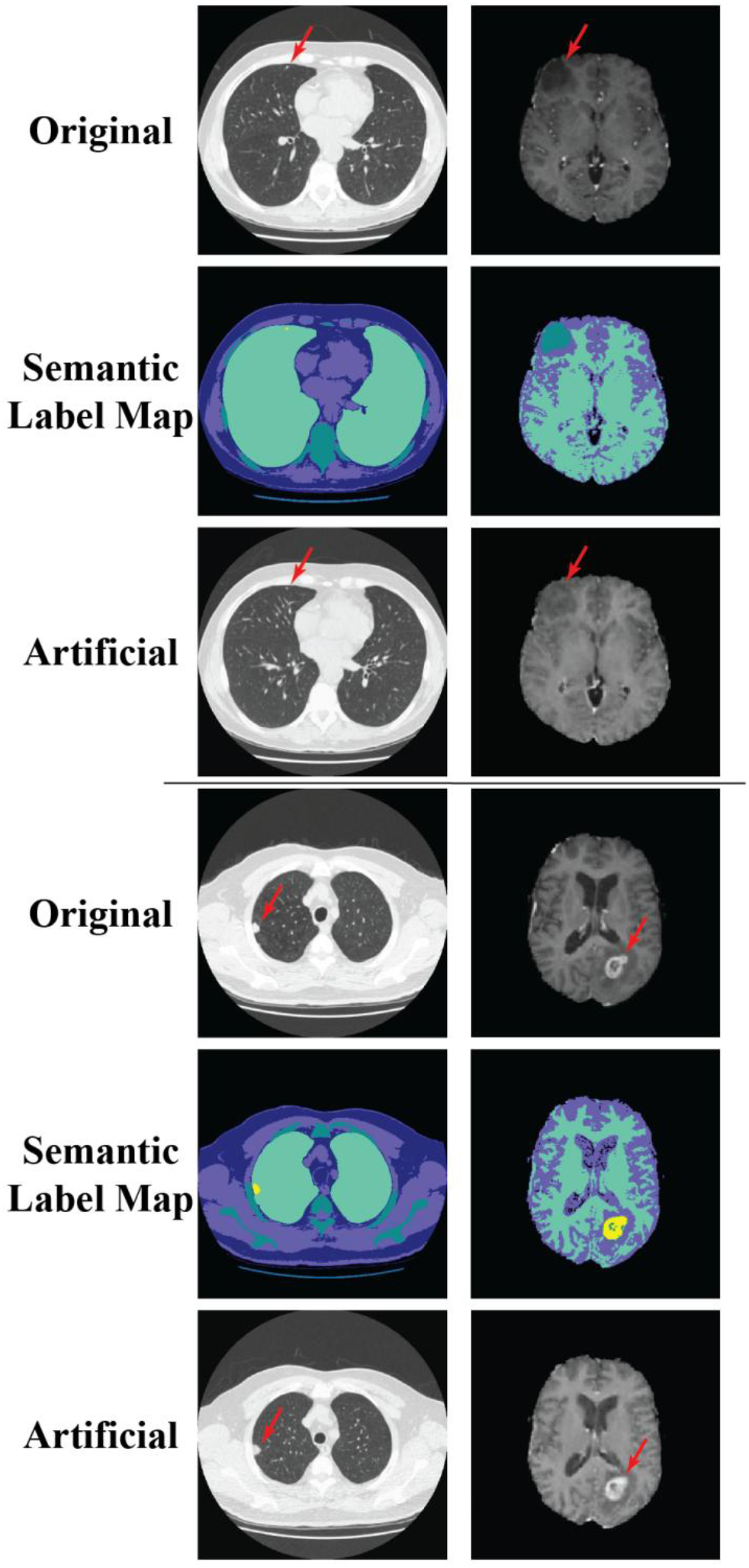
Two CT (left) and MRI (right) slices showing original images along with the corresponding semantic label maps and generated artificial images. The first three rows show a benign nodule (left, CT) and low-grade glioma (right, MRI). The last three rows show a malignant nodule and high-grade glioma.

**Figure 3.**
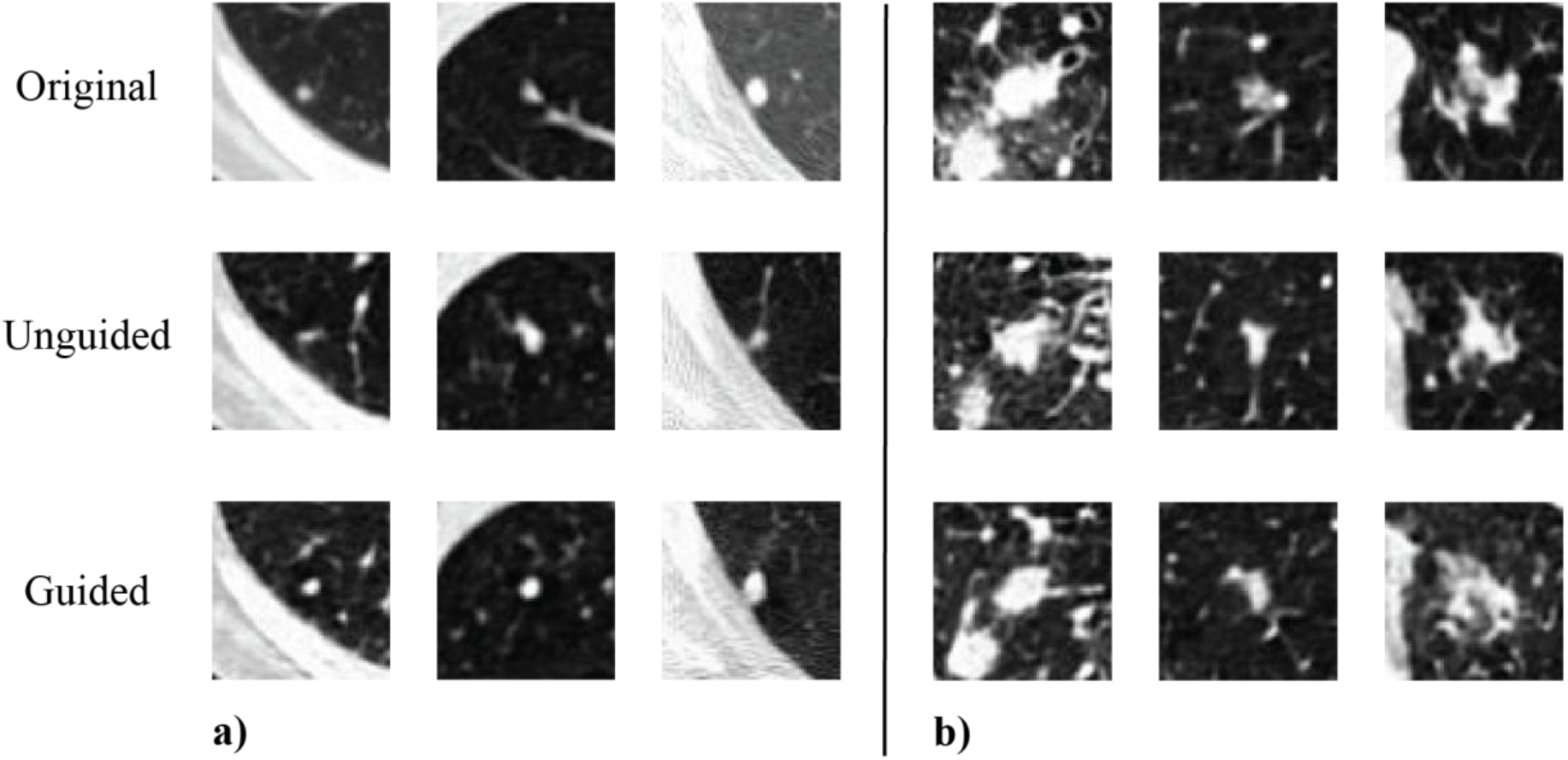
Qualitative representation of the generated images guided by fine features on the LIDC-IDRI dataset: 64x64 cropped areas of interest from the created 2D chest CT artificial images. The first row shows original images, second shows artificial images from the DiFine model without guidance, and the third row shows the artificial images created by DiFine model using independent classifier guidance. a) three benign examples (column 1, 2 and 3) and b) three malignant examples (column 4, 5 and 6).

The independent classifier network trained on the chest CT images achieved a classification accuracy of 72% (0.81 precision and 0.82 recall) on the test set. This represented a ‘baseline’ accuracy based on the original images. The pathologies in images created by the DiFine model without classifier guidance achieved an accuracy of 70%. This was not significantly different than that achieved by images created through traditional GAN-based model with SPADE (69%) (vs unguided, *p =* 0.78). DiFine model guided by the fine features achieved 85.5% accuracy (vs GAN with SPADE, *p* < 0.001, vs unguided, *p* < 0.001; Figure 4). Similarly, in case of the gliomas, differences in area under ROC were significantly higher (81% unguided vs 86% guided (*p <* 0.001)) than the baseline accuracy (Figure 4).

**Figure 4.**
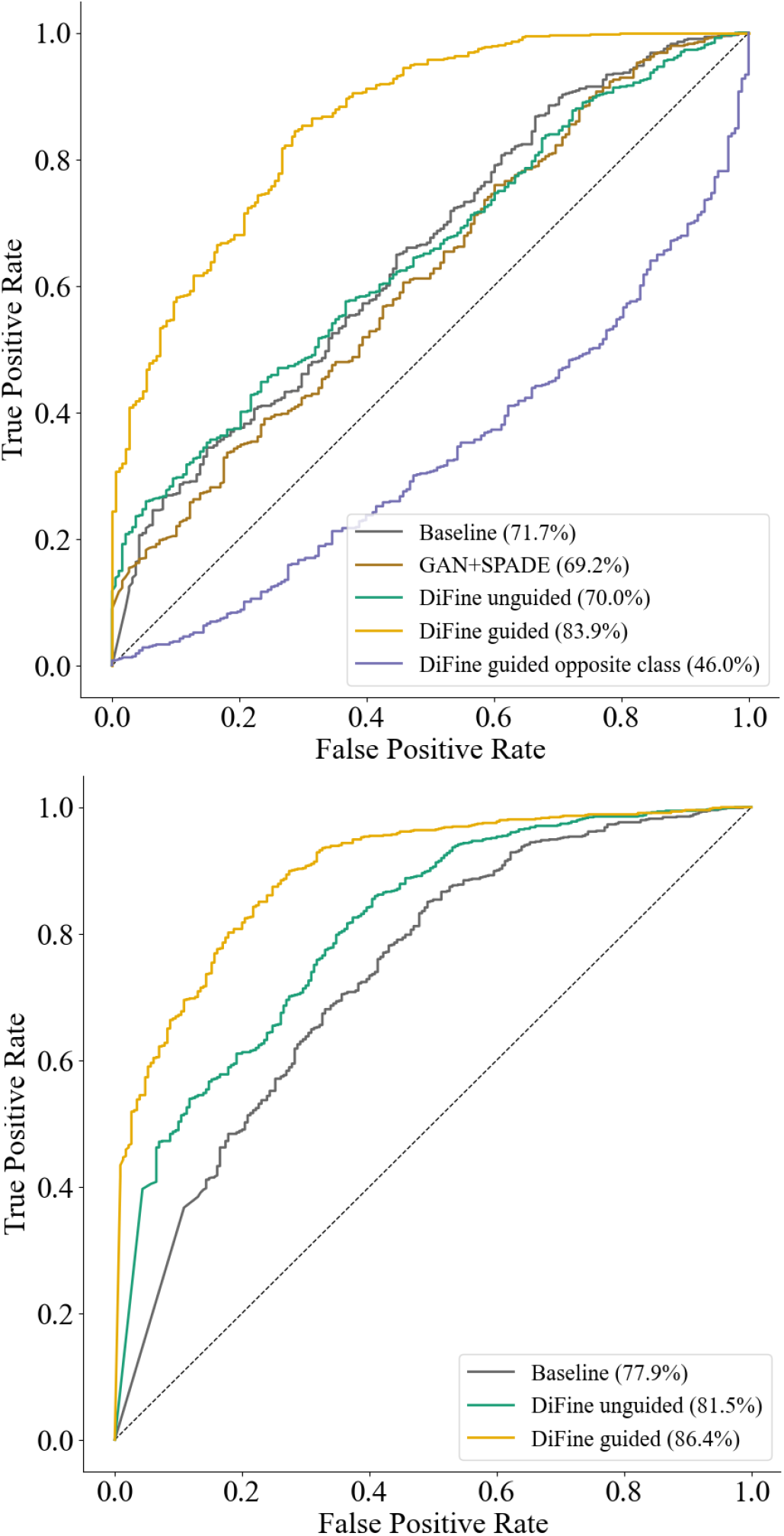
Receiver operating characteristic (ROC) curves of the performance of the classifier on different sets of images. Top: the baseline classification accuracy based on original images from the test set of LIDC-IDRI, on images created by the GAN with SPADE model, on DiFine model without fine-feature guidance, and the DiFine model with independent classifier guidance (trained and tested on both the correct and incorrect “opposite” class). Bottom: Independent classifier performance on images from the BraTS2019 test set.

Figure 4 also shows the ROC curve from our sensitivity analysis (classification of the pathology based on images generated using the opposite class). Sensitivity analysis results are consistent with the main results in that predictive performance was “reverse” (i.e., < 0.5), confirming that the DiFine model applies the type of pathology associated with the target fine features.

## Discussion

We demonstrated the accuracy and feasibility of manipulating fine features of artificial radiographic images, using the feedback from an independent classifier, to reflect different pathological characteristics. This applies across multiple modalities (CT, MRI) pathologies (lung nodules and gliomas), and pathological classification (malignant vs benign, high vs low grade). Traditional GAN models and DiFine without independent classifier fine-feature guidance achieved a classification accuracy comparable to the accuracy of the baseline classification (71.7%). DiFine model with independent classifier fine-feature guidance improved discrimination of pathological designations by almost 15% (to 85.5%). This was consistent for MRI-based glioblastoma and CT-based nodule classification. This highlights the generalizability of DiFine architecture in medical image modalities.

Guidance based on an independent classifier network for diffusion models has been described before ^15^ though the prior work focused on the representation of the entire artificial image. Instead, we focused on an area of interest and processed the classifier feedback in an ‘in-painting’ style. Other prior research focused on improving the quality of finer details in medical images. For example, prior GAN-based models sought to improve edge details by using guidance similar to semantic label maps ^28^. In clinical context, Pinaya et al. ^29^ used a conditional diffusion model to influence characteristics such as sex, age and ventricular volume in T1w MRI artificial brain images. This guidance was implemented in a similar way to Rombach et al. ^13^ In both models, information is concatenated directly with the noisy images. Conceptually, this work is similar to ours in that both used clinical information that can be embedded in the image. However, contrary to our work, neither study used a feature classifier, but used the direct concatenation approach. Thus, in our work, features representing pathology type are learned directly by the classifier as opposed to manually defined gross patient characteristics such as age and sex.

Our model has several limitations. First, the use of a binary mask before in-painting the fine-feature guidance may not be optimal. We used binary masks to prevent any unwanted changes outside of the pathological area. Malignant pathologies are usually larger in size, but the binary mask does not allow the DiFine model to enforce any changes outside of the pathological area. In other words, our architecture does not allow the pathological area to increase in size. Second, we assume that these discriminating features learned by the independent classifier are meaningful, i.e. that they do represent clinically relevant features that indicate the type of pathology. This may or may not be true. We did not seek to “validate” clinical relevance of these learned fine features. Admittedly, this is important for clinical validation of artificial images, but it is inconsequential to the generative model performance. Third, we focused on 2D images and use only one characteristic for fine-feature guidance. Generating 3D images requires substantially higher computational power that is not readily available to us. Nonetheless, creating small 3D medical images using diffusion models is possible ^30^ and we have no reason to believe that this is not the case for our model. Furthermore, our architecture trivially allows training multiple classifiers with different pathological characteristics to provide combined guidance, although we only used a single classifier as the number of annotated pathologies were limited.

Despite its limitations, our work underscores the importance of using independent classifier guidance to accurately represent fine features in generated images and objects, demonstrates feasibility of our architecture across clinical imaging modalities, organs, and pathologies, and highlights its utility in a clinical setting. This can allow effective case simulations for training in a safe environment ^6,31^ or generation of realistic clinical images for training other classification models when the number of training data is limited.

## Acknowledgements/Funding

This work was supported by a ZonMw Innovative Medical Devices Initiative (IMDI) subsidy for the B3CARE project (Dossier number: 10-10400-98-008). The funder had no role in study design, data collection and analysis, decision to publish, or preparation of the manuscript.

Part of this work has been previously included in the PhD thesis of EISH (doi: 10.3990/1.9789036560856).

The authors acknowledge the National Cancer Institute and the Foundation for the National Institutes of Health, and their critical role in the creation of the free publicly available LIDC/IDRI Database used in this study.

## Data availability

The code used in this work can be found in the following repository: https://github.com/UT-RAM-AIM/FineFeature-guidance. The basis of the code is reused from Rombach et al. (2022) and Dhariwal et al. (2021) as mentioned in the text and our code repository. Furthermore, both datasets used, the Lung Image Database Consortium and Image Database Resource Initiative (LIDC-IDRI) and international multimodal Brain Tumor Segmentation Challenge (BraTS2019), are publicly available.

## Author Contributions

All authors contributed to the conception of the work. EISH and XZ reviewed the literature and processed the data. XZ drafted the first and subsequent versions of the main code and EISH contributed to critical revision and drafted data processing code. EISH drafted the first and subsequent versions of the manuscript. FvdH and COT contributed to its intellectual content and to the critical revision of the manuscript. All authors approved the final manuscript for submission and have agreed to be accountable for all aspects of the work.

