## Supplemental Materials and Methods for "It’s All in the Details: Guiding Fine-Feature Characteristics in Artificial Medical Images using Diffusion Models"

### Supplemental information to Materials and Methods

### Network and Guidance Details

In traditional diffusion models, a Gaussian noise is gradually added to the original input image (*forward diffusion*), and the resulting 'noise image' is de-noised recursively to obtain the artificial image (reverse or *backward diffusion*). Traditionally, during *inference*, (i.e. testing) only backward diffusion is used to create a new artificial image. In particular, during the backward diffusion process a deep learning network, usually based on U-Net[1], is used to minimize the noise, producing, in effect, an artificial image ideally identical to the original images.

One of the disadvantages of diffusion models is that training and inference are computationally expensive, especially when large images are used. Latent Diffusion Models (LDMs) [2] were suggested as alternatives to reduce computational cost. In LDMs, model training takes place in a *latent space* to allow embedding of the actual set of images in a lower dimensional space, reducing their size considerably. [2,3]

In their basic form, LDMs create arbitrary artificial images based on the distribution obtained from the actual ones. Instead, it is conceivable that one can *guide* the representation, or features, of the artificial image during the backward diffusion process. In particular, coarse features, e.g., larger anatomical structures, can be guided by *semantic label maps* that represent different objects depicted in the image. These maps are akin to image segmentation masks; each pixel that corresponds to the same object, or feature category, is assigned the same segmentation label. To this end, incorporating so-called SPADE [4] blocks has been shown to be an effective way of guidance by semantic label maps. [5] However, SPADE blocks do not allow guiding the finer features of the individual objects.

#### *Autoencoder Loss Function*

To simplify the autoencoder, we follow one of the implementations of training the autoencoders described by Rombach et al.[6], which contains a reconstruction loss, an adversarial loss, and a KL-divergence regularization loss.

*Denoising Neural Network Structure (UNet) and coarse feature guidance*

To control the coarse features of the artificial image, we use semantic label map guidance. Information of the semantic label map is incorporated by adding SPADE blocks[7] in the diffusion model (see Figure 1 in the main text). SPADE blocks were initially proposed and implemented as an extension to GANs[7]. In this scheme, semantic label maps contain a varying number of labels but consist of at least 2 labels indicating the entire body area and the pathology area. The artificial images adhere to the anatomical constraints imposed by the semantic label map and associated anatomical and gross pathological features. We followed the approach and implementation described by Wang et al..[8]

To better utilize the semantic labels, we replace the GroupNorm normalization layers in the ResNet blocks of the diffusion model's UNet with the specified SPADE normalization layers following Wang et al.[8]. In this scheme, SPADE layers modulate the activations with the semantic labels spatially. Combined with the concatenated channels of the semantic labels in each ResNet block, we formulate a mixed method to implement coarse-feature guidance with the semantic labels, which benefits both inference fidelity and training stability.

*Classifier and Fine-Feature Guidance*

We further guided the model with an in-painting classifier to influence fine-feature representation within an area that contained pathological features. The area of interest (in our context the area where the pathology is evident) is indicated by a specific label in the semantic label map. An independent classifier is trained to distinguish if pathology features are benign or malignant in this area. The guidance from the classifier is implemented in the denoising backward diffusion steps (Figure 1 in the main text) on the areas of interest only if a pathological area is indicated by the semantic label map. This guidance is accompanied by a weight factor or "guidance scale", $s$, which represents the weight applied to the fine-feature guidance from the independent classifier. In other words, a higher value of $s$ will result in more adjustments in the artificial image to present the features of type of pathology, whereas a small value of $s$ will only slightly enforce adjustments. For the LIDC-IDRI and BraTS2019 dataset a guidance scale of s=32 and s=48 is used, respectively.

Before the fine-feature guidance the artificial image is still in a down-sampled state (128x128x3) as the process takes place in the latent space. However, the classifier is trained in the original image space (512x512x1) on the cropped images areas of size 64x64x1. Consequently, the down sampled image is first decoded into the original image space, after which the area of interest is cropped out. To enable fine-feature guidance, we present masked independent classifier guidance concentrated on this area of interest with pathology features. Unlike the previous classifier guidance methods[9], we train the feature classifier on the cropped images of these areas and guide the reverse diffusion process with a mask to avoid unrelated guidance outside the area of interest. To guide the generation process directly in the latent space, we combine the classifier with the pre-trained decoder to calculate the derivatives with respect to the pathology feature in the latent space during inference. Mathematical details are described below in comparison to the work of Dhariwal et al.[9] who applied similar guidance on the full artificial image presentation.

<u>*Mathematical description of inpainting classifier*</u>

The gradients of the derivative of the classifier are used to guide the backward diffusion process (inference). In the original implementation the gradients are used to guide the diffusion process towards a class label (or type of pathology) *y*:

$$\nabla_{x_t} log \boldsymbol{p}_{\Psi}(y|x_t, t)$$

where $x_t$ is a noisy image during the inference process of the diffusion model and $\boldsymbol{p}_{\Psi}(y|x_t, t)$ represents the classifier at time step *t*. In contrast to the original implementation, we used local in-painting style guidance instead of using the whole gradient. Since we used a model that works in the latent space, the noisy latent codes $z_t$ need to be decoded into the original pixel space using the trained decoder $D$, resulting in $x_t = D(z_t)$. We further used a cropped area of interest, in case of chest CT of 64x64x1, to focus on the location where we want to emphasize the fine-feature influence. The locally guiding classifier in our case can be described as:

$$\boldsymbol{p}_{\Psi}(y|x_t, t) = \boldsymbol{p}_{\Psi}(y|Crop(D(z_t)), t)$$

To prevent undesirable artefacts, we make additional use of a binary image mask $\mathbf{M}$. This binary image mask is generated around the region of interest, the pathological area, using a max-pooling operation in the latent space, after which the guidance is only implemented on the non-masked region of the cropped image. This results in a masked gradient in the latent space:

$$(1-\boldsymbol{M})\nabla_{z_t} log \boldsymbol{p}_{\Psi}(y|Crop(D(z_t)),t)$$

The influence of the classifier on the artificial image region of interest can be emphasized by increasing a guidance scale $s$ of the gradient, that is $s\cdot\nabla_{z_t}\log \boldsymbol{p}_{\Psi}(y|Crop\big(D(z_t)\big),t)$. With a larger $s$, there is more focus on the modes of the classifier, consequently emphasizing characteristics of the desired class type in the artificial image area.

**Data preprocessing**

For both datasets, we give a brief overview of processing steps, and the detailed specific implementation can be found in the code available at: https://github.com/UT-RAM-AIM/FineFeature-guidance.

From the Lung Image Database Consortium (LIDC) and Image Database Resource Initiative (IDRI) we used 805 low-dose chest CT scans[10,11]. The 2D 512x512 image slices as well as the corresponding semantic label maps were obtained in a similar method as in[12]. The semantic label maps define 5 labels: body area, soft tissue area, high-dense tissue area, lung area, and lung nodule area (see Table 1 in the main text).

International multimodal Brain Tumor Segmentation Challenge (BraTS2019) dataset[13–17] contains 259 cases with high- and low-grade glioblastoma. All multimodal scans consist of T1-weighted, T1 contrast enhanced (T1ce), T2-weighted, and T2 Fluid Attenuated Inversion Recovery (T2-FLAIR), obtained over various institutions, scanners and with different protocols. For this study, we only use one MRI sequence, T1ce. This sequence provides improved visualization of vascularization, which helps identify masses[18,19]. Standard image processing techniques were used to obtain 240x240 2-dimensional images:

histogram normalization and possible resizing. The semantic label maps were created with 5 labels: background, white matter, gray matter, necrotic tumor core and GD-enhancing tumor. The first three were obtained using Otsu's multi-thresholding method. The original dataset was accompanied by segmentations of the glioma area.
